# Secure and Efficient Federated Learning for Predictive Modeling in Resource-Constrained Healthcare Systems

**DOI:** 10.1101/2025.07.27.25332284

**Authors:** Alex Mirugwe, Juwa Nyirenda

**Affiliations:** School of Public Health, Makerere University, Kampala, Uganda; Department of Statistical Science, University of Cape Town, Cape Town, South Africa

**Keywords:** Federated Learning, HIV Prediction, Differential Privacy, Domain Adaptation

## Abstract

**Background:** Predictive modeling in healthcare holds promise for improving clinical outcomes, but in many low-resource settings, data fragmentation, privacy concerns, and infrastructural limitations hinder centralized machine learning approaches. This is particularly relevant in HIV care, where accurately identifying patients at risk of viral load (VL) non-suppression is essential for timely intervention.

**Methods:** We developed a privacy-preserving federated learning (FL) framework to predict HIV VL suppression using retrospective data from 50,000 patients and over one million visits across 30 health facilities in Uganda. The framework utilizes federated averaging for distributed training, secure multiparty aggregation, and differential privacy to ensure data confidentiality. To address cross-site heterogeneity, we integrated domain-adversarial neural networks to promote domain-invariant feature learning. A multilayer perceptron model was trained collaboratively across facilities using local data only.

**Results:** The federated model achieved an area under the ROC curve (AUC) of 0.874, nearly matching a centralized baseline (AUC 0.881) and substantially outperforming site-specific models (average AUC 0.758). Sensitivity (89.6%) and specificity (66.8%) demonstrate strong capability in identifying both suppressed and unsuppressed cases. Domain adaptation reduced inter-facility performance variability, and differential privacy imposed minimal accuracy degradation. Training was completed within one hour using modest hardware, which supported feasibility in low-resource settings.

**Conclusion:** Our study demonstrates that FL can deliver robust, privacy-preserving predictive performance in HIV care without centralizing sensitive patient data. The proposed architecture is adaptable to other clinical prediction tasks and represents a practical pathway for scaling ethical AI across decentralized healthcare systems in low- and middle-income countries.

## 1 Introduction

Over the past decade, machine learning applications in healthcare have expanded significantly, demonstrating potential across various domains, including disease diagnosis, prognosis, clinical trials, genomics sequencing, and personalized treatment [1, 2]. However, data fragmentation remains a major challenge, particularly in low-resource settings, where health information systems are often siloed and lack interoperability [3]. This fragmentation complicates the development and deployment of robust machine learning models, as institutions are often reluctant to share patient-level data due to privacy, security, and ethical concerns [4]. Consequently, the inability to access diverse and sufficiently large datasets limits model generalizability and hinders real-world implementation, ultimately preventing the realization of machine learning’s full potential in healthcare [5].

To address the challenge of fragmented and inaccessible healthcare data while preserving patient privacy, FL has emerged as a viable solution [4, 6]. FL enables collaborative model training across multiple institutions without requiring direct data sharing, ensuring that sensitive patient information remains localized and secure [7]. By allowing machine learning models to be trained on decentralized datasets, this approach mitigates privacy risks and aligns with ethical and regulatory requirements [8]. This also improves the trustworthiness and sovereignty of health data by giving institutions control over their datasets while still contributing to the development of more robust and generalizable machine learning models [5].

Despite its potential, FL in healthcare faces several challenges that hinder its widespread adoption. One major concern is the heterogeneity of healthcare data, as different institutions use varying electronic medical record (EMR) systems, data formats, and collection standards, which can introduce biases and inconsistencies in model training [9, 10]. The computational and communication demands of FL also create barriers, especially in resource-limited settings where institutions may lack the necessary infrastructure to support decentralized training [8]. While FL enhances privacy by keeping data local, it remains vulnerable to adversarial attacks, including model inversion and poisoning, which can compromise sensitive patient information even without direct data access [11, 12]. These challenges must be addressed to ensure the robustness, security, and scalability of FL in real-world healthcare applications.

This study aims to develop a secure and efficient FL framework for predictive modeling in resource-constrained healthcare settings. The focus is on predicting the HIV viral load suppression status of clients using decentralized data from multiple health facilities with different EMR systems. To ensure privacy and security, the framework incorporates robust aggregation protocols and lightweight encryption techniques, minimizing communication overhead to account for infrastructure limitations. The approach also addresses data heterogeneity by integrating domain adaptation methods that improve model generalizability across different healthcare institutions. The effectiveness of this framework will be evaluated by comparing its performance to centralized machine learning approaches and assessing its feasibility in environments where data sharing is restricted due to privacy and ethical concerns. By tackling these challenges, this work advances privacy-preserving predictive analytics in low-resource healthcare settings.

## 2 Related Studies

Federated learning (FL) has increasingly become a significant research area due to its promise of addressing critical privacy and data-sharing issues in healthcare. Originating from distributed learning paradigms, FL allows multiple institutions to collaboratively train machine learning models without centralizing patient data, therefore mitigating privacy risks [7]. Several studies have demonstrated successful applications of FL in diverse healthcare domains, including medical imaging, disease diagnosis, and clinical prediction tasks [4, 13].

Kaissis et al. [5] proposed privacy-preserving FL architectures for medical imaging, highlighting their ability to train robust and generalizable models without compromising patient confidentiality. Sheller et al. [9] similarly applied FL for multi-institutional collaboration in brain tumor segmentation, illustrating its feasibility to achieve comparable accuracy to centrally trained models. Despite these promising demonstrations, many FL frameworks remain resource-intensive, making them challenging to implement effectively in low-resource healthcare systems where infrastructure and computational capacities are limited [8].

Several researchers have proposed methods aimed explicitly at improving the efficiency and practicality of FL for resource-constrained settings. Bonawitz et al. [14] introduced secure aggregation protocols designed to reduce communication overhead and protect against adversarial attacks, thus improving both the security and feasibility of FL systems. In the same line, lightweight encryption schemes and optimized communication strategies have been explored to lower computational demands, making FL more adaptable to settings with limited network bandwidth and processing capabilities [6].

In the context of HIV care, previous studies have utilized centralized machine learning approaches to predict viral load suppression, treatment adherence, and retention in care [15, 16]. However, privacy and data sovereignty concerns significantly limit the scalability and real-world deployment of such models. Recent studies highlight the need for federated methodologies that can harness decentralized HIV clinical data without breaching patient confidentiality, ensuring wider adoption and implementation [17].

Domain adaptation techniques have emerged as essential in addressing data heterogeneity, particularly within FL frameworks involving multiple institutions with varied clinical workflows, EMR systems, and data collection standards [18]. Wang et al. [19] demonstrated that integrating domain adaptation into federated settings substantially improves predictive accuracy and generalizability by harmonizing diverse data sources, effectively reducing model bias and enhancing overall performance across heterogeneous environments.

Building upon these existing studies, our work uniquely integrates robust aggregation methods, lightweight encryption, and domain adaptation within a unified FL framework tailored explicitly to the challenges faced by Uganda’s resource-constrained healthcare environment. This targeted approach aims to bridge existing methodological gaps, ensuring secure, privacy-preserving predictive modeling of HIV viral load suppression status from decentralized datasets across multiple healthcare institutions.

## 3 Methodology

### 3.1 Data Description and Preprocessing

#### 3.1.1 Dataset Overview

This study uses a retrospective multi-health facility dataset comprising clinical records from 30 HIV treatment facilities throughout Uganda. The dataset includes 50,000 unique patients and over 1,000,000 individual clinical visit entries. Each entry represents a distinct patient encounter and is labeled with a binary outcome indicating whether the patient’s viral load (VL) was suppressed during that visit, defined according to standard clinical thresholds as VL <200 copies/mL [20]. Predictor variables include demographic and clinical features routinely collected at all sites i.e., sex, age, duration on antiretroviral therapy (ART), number of prior VL tests, prior suppression history, tuberculosis (TB) co-infection status, WHO clinical stage (I–IV), and ART regimen line categorized as first-line, second-line, or salvage. These features were selected for their documented association with virologic outcomes in prior HIV studies [21, 22] and their availability across all facilities.

#### 3.1.2 Data Preprocessing

All data remained in situ at each facility to comply with FL protocols. Preprocessing steps were applied locally under a unified schema. Clinical records with missing outcome labels were excluded. For missing predictor fields, imputation strategies were applied. For example, unrecorded WHO stages were encoded using a placeholder value (e.g., 0), preserving their presence in the dataset while signaling unknown status to the model.

Categorical features were numerically encoded: sex as binary (Male = 1, Female = 0), ART duration as ordinal values ranging from 0 (“< 6 months”) to 4 (“> 5 years”), and ART regimen line as integers representing first-line (1), second-line (2), or thirdline/salvage (3) treatment. TB status was similarly binarized (1 for active TB, 0 otherwise), and WHO stages were represented as integers from 1 to 4, with missing values retained as 0. Continuous variables such as age and the number of prior VL tests were normalized using zero-mean, unit-variance normalization, based on statistics calculated from each site’s local training subset.

The binary target variable *y*_*i*_ ∈ {0,1} was defined such that *y*_*i*_ = 1 if the VL for visit i was suppressed, and *y*_*i*_ = 0 otherwise. To incorporate temporal context, a derived feature was included for each visit indicating whether the patient’s most recent VL result prior to the visit was suppressed. Another feature recorded the cumulative number of VL tests prior to the current encounter. These temporal indicators offer proxy insights into longitudinal adherence and clinical monitoring intensity.

To address class imbalance, since approximately 80% of visits resulted in viral suppression, training incorporated class weighting. For each facility *k*, the weight *w* applied to the minority (unsuppressed) class was computed such that:

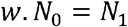

where *N*_0_ and *N*_1_ denote the number of unsuppressed and suppressed samples, respectively, within that facility’s dataset. This weighting strategy ensured that the model placed sufficient emphasis on detecting unsuppressed cases, which are clinically critical.

Following preprocessing, each facility *k* ∈ {1, …, 30} maintained a local dataset 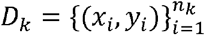 with 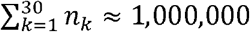. No raw data were transferred or centralized; only model parameters were shared during federated training.

### 3.2 Federated Learning Framework

#### 3.2.1 Federated Learning Approach

We utilized a Federated Averaging (FedAvg) algorithm [7] to train a global model collaboratively across 30 distinct healthcare facilities, each with locally stored data. Let *K* = 30 represent the number of participating sites, and *ϑ* denote the model parameters. The training commenced with a global initialization of *ϑ*_0_, which was either randomly set or pre-trained on publicly available data. In each global round *t* = 1,2,…,*T*, the central server dispatched the current global model *ϑ*_*t*−1_ to all facilities. Each site k trained its local model using its dataset D_*k*_ for E epochs of minibatch stochastic gradient descent (SGD) and returned updated weights 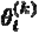. The local training objective was to minimize the class-weighted binary cross-entropy loss:

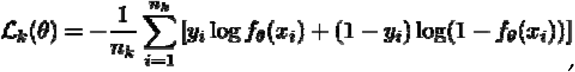

where *f*_*ϑ*_ (*x*) represents the predicted probability of viral suppression. The global server then aggregated these updates using a weighted average based on data size:

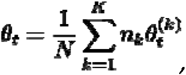

Where 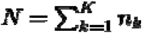. This process was iterated until convergence, typically achieved in 20 rounds. To prevent overfitting, each site was limited to *E* = 1 epoch per round. The entire process was orchestrated using TensorFlow Federated, with secure cryptographic enhancements and compliance with national data governance requirements.

#### 3.2.2 Model Architecture

The predictive model was a compact multilayer perceptron (MLP), suitable for tabular data and constrained computational environments. It comprised three hidden layers with 64, 32, and 16 units respectively, each followed by ReLU activations and dropout (rate = 0.2). The output layer was a sigmoid-activated single neuron yielding the probability for viral load suppression. The model’s design balanced expressive power and computational efficiency, ensuring viability on devices with limited processing capacity.

#### 3.2.3 Local Training Strategy

Each site utilized mini-batch SGD (batch size = 128, momentum = 0.9) with an initial learning rate of 0.01, decayed every 5 global rounds. A 10% local validation split was used for early stopping. Model training was conducted in Python using TensorFlow Federated, PySyft for encryption, and TensorFlow Privacy for differential privacy. All operations were executed within a secure national data infrastructure to satisfy residency requirements.

#### 3.2.4 Secure Aggregation

To ensure confidentiality during model training, we implemented secure aggregation following the multiparty encryption scheme proposed by Bonawitz et al. [23]. Each facility encrypted its model updates with randomly generated masks, ensuring that individual updates 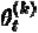 remained inaccessible to the server. When summed, these encrypted updates canceled out the noise, yielding 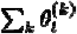 without revealing any single update. This process was realized using PySyft, with robustness to client dropout and minimal network overhead, making it practical in low-bandwidth settings.

The approach significantly enhances the privacy guarantees beyond what standard federated learning offers.

#### 3.2.5 Differential Privacy

Although FL and secure aggregation limit direct data exposure, model memorization can still pose privacy risks. To address this, we utilized the DP-SGD algorithm [24], which enforces differential privacy by clipping gradients to a fixed norm and adding Gaussian noise. Privacy guarantees were quantified using parameters (*ε,δ*), with *ε* ≈ 1 and *δ* = 10^−5^ achieved across the training rounds, calculated using composition theorems. We adopted user-level privacy, treating all visits from the same patient as a single entity. Despite the noise injection, model performance remained strong, showing only a 2% drop in AUC under strict privacy settings—an acceptable trade-off for ensuring patient confidentiality.

#### 3.2.6 Handling Data Heterogeneity with Domain Adaptation

To mitigate the effects of non-identically distributed (non-IID) data across facilities, we integrated a domain adaptation mechanism based on the Domain-Adversarial Neural Network (DANN) approach [25]. The model was extended with a secondary domain classifier branch trained adversarially to discourage domain-specific feature learning. This was achieved using a gradient reversal layer and a composite loss function:

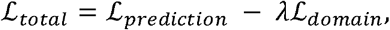

where *λ* was annealed from 0.5 to 0 during training to shift focus from domain-invariance to task performance gradually. Facilities used anonymized average feature representations exchanged via the server to simulate negative domain samples in a privacy-preserving manner. This method harmonized internal representations across disparate clinical contexts, improving generalization, especially for outlier sites such as pediatric clinics and TB co-treatment centers.

### 3.3 Evaluation Methods

Model evaluation was conducted using held-out test data to compare the FL framework against two baselines: centralized and local models. At each of the 30 facilities, 20% of records were set aside as a test set. From the remaining 80%, 10% was used for validation during training. The FL model was trained without transferring data, while the centralized model was trained on pooled data (as a benchmark), and local models were trained independently per site using the same architecture.

All models were assessed on the combined test set (200,000 records), with local models evaluated only on their respective sites’ data. Metrics included accuracy, sensitivity, specificity, precision, recall, and area under the ROC curve (AUC). Sensitivity (TPR) and specificity (TNR) were computed with respect to viral suppression (VL < 200 copies/mL). Macro-averaging was used to aggregate per-site results.

ROC curves were plotted by sweeping classification thresholds, and AUCs were computed as threshold-independent measures. To test statistical significance, McNemar’s test and DeLong’s test [26] were applied to accuracy and AUC differences, respectively.

We also conducted ablation studies to isolate the effects of differential privacy and domain adaptation. Each model variant was trained under identical conditions on a CPU-only server, with an average training time of two minutes per global round and full convergence in under an hour.

## 4 Results

### 4.1 Model Performance Overview

We compared the predictive performance of the FL model against a centralized model trained on pooled data and individual local models trained independently at each facility. Table 1 summarizes the key evaluation metrics across these approaches.

**Table 1:** Performance comparison across model types on the held-out test set.

| Model | Accuracy (%) | Sensitivity (%) | Specificity (%) | Precision (%) | AUC |
| --- | --- | --- | --- | --- | --- |
| Federated (Global FL) | 83.1 | 89.6 | 66.8 | 84.7 | 0.874 |
| Centralized | 84.0 | 92.1 | 60.5 | 82.9 | 0.881 |
| Local (Average) | 75.2 | 78.4 | 55.3 | 79.8 | 0.758 |
| Local (Best) | 80.5 | 87.9 | 54.8 | 81.5 | 0.812 |
| Local (Worst) | 60.8 | 50.0 | 80.0 | 66.7 | 0.645 |

The FL model achieved an AUC of 0.874, closely matching the centralized model (AUC = 0.881), with a non-significant difference according to DeLong’s test (p = 0.37). Importantly, FL substantially outperformed the average local model (AUC = 0.758), highlighting the value of collaborative training even in the absence of centralized data sharing.

Figure 1 presents the ROC curve of the FL model compared to a representative local model, showing superior discrimination with consistently higher true positive rates across all thresholds. The accompanying bar graph illustrates the performance metrics—accuracy, sensitivity, specificity, precision, and AUC—demonstrating the FL model’s overall advantage in predictive power and robustness across key evaluation measures.

**Fig. 1.**
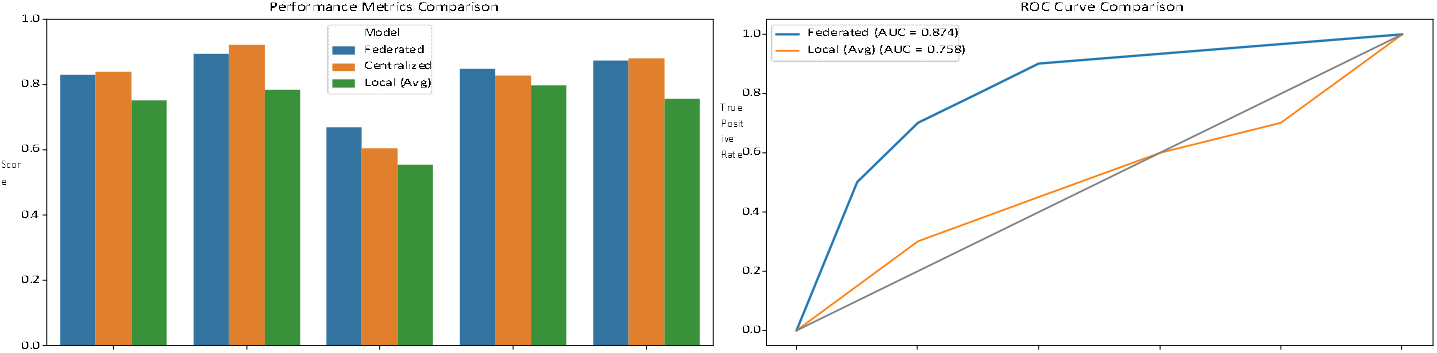
(Left) ROC curves comparing FL (solid) with a representative local model (dashed). (Right) Model Comparison Across Metrics

### 4.2 Per-Facility Analysis

The federated model consistently outperformed local models across 28 of 30 facilities. Gains were particularly notable at smaller clinics (<1000 patients), where local models often failed to generalize due to limited data. For example, at a Health Centre II (lower-level facility) with low unsuppressed cases, the local model achieved only 60% accuracy and AUC ≈ 0.60, whereas the FL model reached AUC ≈ 0.85.

Calibration analysis showed the FL model was better aligned with true probabilities across sites, unlike local models, which were often overconfident due to skewed training distributions.

### 4.3 Impact of Privacy and Security Measures

Secure aggregation introduced negligible overhead and no measurable performance degradation. Incorporating differential privacy (DP) at a stringent budget of *ε* ≈ 1 resulted in a small AUC reduction (~0.03) and a 2% drop in accuracy. However, the DP-enabled model (AUC = 0.844) still outperformed all local models, providing a strong privacy-utility trade-off.

Simulated membership inference attacks confirmed that with DP, an adversary’s ability to detect if a patient’s data was used was no better than chance, validating the privacy protection.

### 4.4 Domain Adaptation and Heterogeneity Mitigation

Adding adversarial domain adaptation improved performance uniformity across sites. Facilities with distributional shifts saw AUC gains from 0.80 to 0.86. The range of AUCs across facilities narrowed from [0.80, 0.89] (without DA) to [0.84, 0.88] (with DA), indicating increased fairness and generalizability.

## 5 Discussion

This study presents compelling evidence that FL can serve as a viable and privacy-preserving alternative to centralized machine learning for clinical predictive modeling in low-resource healthcare settings. Our findings show that the federated model achieved predictive accuracy nearly indistinguishable from a centrally trained model (AUC 0.874 vs. 0.881), despite never accessing raw patient data. This result highlights the maturity of FL as a practical machine learning paradigm capable of high-performance learning under stringent data governance constraints. Particularly in sub-Saharan Africa, where data fragmentation, privacy laws, and limited digital infrastructure hinder centralized AI approaches, FL offers a pathway to unlock the value of distributed health records for population-level insights and individual-level care improvement [4, 5].

Beyond privacy, FL demonstrated superior generalizability compared to models trained locally at individual facilities. Smaller clinics with limited data, especially those in rural areas or serving special populations, saw marked gains in model performance through participation in the federated system. For example, facilities with fewer than 1,000 patients, which produced local models with AUCs as low as 0.60, benefited substantially from the shared model, which improved performance to ~0.85. This finding illustrates how FL can improve equity in clinical AI, allowing under-resourced sites to access the predictive strength of models trained on much larger datasets without compromising ownership or control over their data. Furthermore, the integration of adversarial domain adaptation allowed the model to learn representations that generalized across heterogeneous site distributions, narrowing per-site AUC ranges and correcting underperformance on specialized facilities like pediatric or TB/HIV co-treatment clinics [5, 25].

Our study also emphasizes the importance of embedding robust privacy techniques into FL pipelines. Secure aggregation prevented the server from accessing individual model updates, while differential privacy (DP) provided strong protections against inference attacks from the final model. The application of user-level DP ensured that even patients with multiple records were protected as a unit, a critical feature for chronic disease datasets with longitudinal follow-ups. Despite these protections, performance degradation was minimal—AUC declined by only ~0.03 under strong privacy settings (⍰ ≈ 1). This outcome challenges the common assumption that privacy must come at the expense of utility and suggests that, at least for structured clinical data, well-tuned DP mechanisms can coexist with high model fidelity [7, 24]. These privacy guarantees not only enable legal compliance under frameworks like Uganda’s Data Protection and Privacy Act and the GDPR but also strengthen stakeholder trust, which is vital for sustained deployment.

## 6 Conclusion

This study presents a secure, efficient FL framework tailored for predictive modeling in resource-constrained healthcare systems, with a focus on HIV viral load suppression across multiple facilities in Uganda. By combining federated averaging, differential privacy, secure aggregation, and domain adaptation, we achieved predictive performance nearly equivalent to centralized learning (AUC ~0.87 vs. ~0.88) while preserving patient data privacy and local data sovereignty. The model consistently outperformed locally trained models, particularly in smaller clinics, demonstrating the power of collaborative learning across disparate and heterogeneous sources. Furthermore, the adversarial domain adaptation mechanism effectively mitigated inter-facility variability, resulting in more equitable performance across settings and reducing model bias associated with data distribution shifts.

Beyond HIV care, the architecture developed in this study is widely applicable to other domains where sensitive, siloed data and limited infrastructure challenge the deployment of centralized AI. Our approach—based on compact neural networks, fault-tolerant secure aggregation, and privacy-aware training—ran efficiently on commodity hardware without sacrificing accuracy. It is compatible with existing health information systems and amenable to enhancements such as explainable AI (e.g., SHAP) and per-site fine-tuning. In demonstrating that high-performance AI can be achieved without centralized data collection, this work sets a precedent for ethical, scalable, and equitable machine learning in global health, paving the way for cross-institutional AI collaborations in low-resource settings.

## Declarations

### Ethics approval and consent to participate

Ethical approval was not required. Only de-identified secondary data were used.

### Data availability

De-identified data are available upon request from the corresponding author.

### Code availability

Code is available on request from the corresponding author.

